# Pathway for Enhanced Recovery after Spinal Surgery-A Systematic Review of Evidence for use of Individual Components

**DOI:** 10.1101/2020.08.16.20175943

**Authors:** Ana Licina, Andrew Silvers, Harry Laughlin, Jeremy Russell, Crispin Wan

## Abstract

**Background:** Enhanced recovery in spinal surgery (ERSS) has shown promising improvements in clinical and economical outcomes. We have proposed an ERSS pathway based on societal recommendations and qualitative reviews. We aimed to delineate the clinical efficacy of individual pathway components in ERSS through a systematic narrative review.

**Methods:** We included systematic reviews and meta-analysis, randomized controlled trials, non-randomized controlled studies, and observational studies in adults and pediatric patients evaluating any one of the 22 pre-defined care components. Our primary outcomes included all-cause mortality, morbidity outcomes (e.g., pulmonary, cardiac, renal, surgical complications), patient-reported outcomes and experiences (e.g., pain, quality of care experience), and health services outcomes (e.g., length of stay and costs). We searched the following databases (1990 onwards)

MEDLINE, EMBASE, and Cochrane Library (Cochrane Database of Systematic Reviews and CENTRAL). Two reviewers independently screened all citations, full-text articles, and abstracted data. A narrative synthesis was provided. Where applicable, we constructed Evidence Profile (EP) tables for each individual element. Due to clinical and methodological heterogeneity, we did not conduct a meta-analyses. Confidence in cumulative evidence for each component of the pathway was classified according to the GRADE system.

**Results:** We identified 5423 relevant studies excluding duplicates as relating to the 22 pre-defined components of enhanced recovery in spinal surgery. We included 664 studies in the systematic review. We found specific evidence within the context of spinal surgery for 14/22 proposed components. Evidence was summarized in EP tables for 12/22 components. We performed thematic synthesis without EP for 6/22 elements. We identified appropriate societal guidelines for the remainder of the components.

**Discussion:** We identified the following components with high quality of evidence as per GRADE system: pre-emptive analgesia, peri-operative blood conservation (antifibrinolytic use), surgical site preparation and antibiotic prophylaxis. There was moderate level of evidence for implementation of prehabilitation, minimally invasive surgery, multimodal perioperative analgesia, intravenous lignocaine and ketamine use as well as early mobilization. This review allows for the first formalized evidence-based unified protocol in the field of ERSS.

Further studies validating the multimodal ERSS framework are essential to guide the future evolution of care in patients undergoing spinal surgery.

## Background

Enhanced recovery after surgery (ERAS) programs have demonstrated improvements in patient outcomes encompassing superior recovery, improved functional measures, lower morbidity, decreased length of stay with healthcare cost savings (1) (2). International disease burden of spinal conditions is high (3). Between 2004 to 2015, volume of elective lumbar fusion increased by 62.3 percent in the United States. There was an accompanying increase in healthcare hospital costs due to lumbar fusion admissions, with a 177 percent increase during the 12-year study period (4). There is a need to apply lessons learned from enhanced recovery programs in other surgical specialties to surgery of the spine (5). Limited enhanced recovery pathways have been applied to spinal surgery, with a uniform finding of decreased length of stay across reported studies (6–9). There was a notable decrease in the adverse events during hospital stay (9,10).

Prior narrative qualitative reviews have delineated recommendations for the incorporation of individual components into an enhanced recovery after spinal surgery (ERSS) program. Several critical components of enhanced recovery in spinal surgery have been identified. These include: provision of comprehensive perioperative nutrition, multimodal analgesia, minimally invasive surgery where clinically feasible and early mobilization (5). The inclusion of various technical elements in the individual programs is often based on the perceived clinical benefit. (11). Individual ERSS programs differ substantially (12). There is gap in the knowledge with regards to defining the components of ERSS. Our group of authors have identified and proposed the first comprehensive program of Enhanced Recovery in Spinal Surgery (*Table 1*)(11). We defined the individual components based on the enhanced recovery protocols in other surgical subspecialties and prior qualitative reviews of ERAS in spinal surgery. (1,12–16) (17–20). Although program components have been defined, evidence based clinical utility of individual components has not been systematically reviewed.

**Table 1.** Components of enhanced recovery in spinal surgery, grouped according to perioperative stage of care

| *Preadmission period* | *Intraoperative period* | *Postoperative period* |
| --- | --- | --- |
| 1. Preadmission information, education and counseling | 9. Prevention of nausea and vomiting | 17. Thromboprophylaxis |
| 2. Risk assessment, preoperative optimization, including lifestyle factor modification | 10. Surgical site preparation and antimicrobial prophylaxis<br>10.1 Surgical site preparation<br>10.2 Antimicrobial prophylaxis | 18. Urinary drainage |
| 2.1 Pre-operative risk stratification | 11. Local anaesthetic infiltration | 19. Postoperative nutrition and fluid management |
| 2.2 Preoperative assessment and optimization | 12. Standard anaesthetic protocol | 20. Postoperative glycemic control |
| 2.3 Alcohol use | 13. Surgical access (open and minimally invasive spinal surgery, including robotic surgery) | 21. Early mobilization |
| 2.4 Tobacco use | 14. Maintenance of normothermia | *Quality of care measures* |
| 3. Prehabilitation | 15. Intraoperative fluid and electrolyte therapy | 22. Audit |
| 4. Preoperative nutritional care | 16. Perioperative analgesia |  |
| 4.1 Nutritional assessment and screening |  |  |
| 4.2 Perioperative immuno-nutrition |  |  |
| 5. Management of anaemia |  |  |
| 6. Perioperative blood conservation strategies |  |  |
| *Preoperative period* |  |  |
| 7. Preoperative fasting and carbohydrate loading |  |  |
| 8. Preemptive analgesia |  |  |

The aim of this study was to systematically evaluate pre-defined individual components of an ERSS pathway (program). These components together form the basis of a first comprehensive enhanced recovery pathway in spinal surgery (ERSS). We plan to create an evidence-based assessment of the available literature for each pre-defined component of an ERSS program(21). Delineating and formulating the evidence base for each component, would strengthen the quality of ERSS programs. Consistency with regards to best multidimensional practice in ERSS, would allow for standardization of care pathways. Greater standardization of care pathways results in improved external validity across proposed comparative research.

## Methods

This systematic review has been performed according the methodological standards for complex reviews (22) (23–28) (29). Our findings have been reported according to the standards for the Preferred Reporting Items for Systematic Reviews and Meta-Analysis (30). Protocol for this review was prospectively registered with the International Register of Systematic Reviews identification number CRD42019135289 (31). The authors identified the essential components of enhanced recovery within the area of spinal surgery. The authors performed this process by reviewing the current externally validated enhanced recovery protocols as recommended by the ERAS Society. We identified and applied the relevant components to the field of spinal surgery (13) (1, 14, 15) (12, 32) (5). We have published this work through a peer reviewed protocol dissemination(11).

### Eligibility criteria

We included adult and paediatric patients undergoing spinal surgical procedure on any anatomical site including cervical (anterior or posterior cervical decompression and fusion), thoracic (e.g., thoracic decompression and fusion), lumbar (e.g., lumbar decompression and fusion, lumbar laminectomy, sacral or any one combination of these). We included systematic reviews and meta-analysis, randomized controlled trials, non-randomized controlled studies, and observational studies (e.g., cohort studies, case-control studies, cross-sectional studies, and case series). We included human data studies published in the English language after 1990.

The interventions of interest have been classified in 5 perioperative pillars: preadmission period, preoperative period, intraoperative period, postoperative period, and audit and compliance processes (*Table 1)*. We reviewed the evidence with regards to each component studied independently or in any one combination (33). Our full review eligibility criteria are listed in *Table 2*.

**Table 2.** Review eligibility criteria

| *Study characteristic* | *Inclusion criteria* | *Exclusion criteria* |
| --- | --- | --- |
| Patient population | Adults undergoing spinal surgical procedures;<br>Paediatric population undergoing spinal surgical procedures; | Patients undergoing non-surgical management of spinal conditions;<br>Spinal trauma patients; |
| Intervention-treatment | Twenty-two pre-defined components of an ERSS pathway (as outlined in Table 1) alone or in combination with another component ;<br>Other proposed ERSS pathways incorporating one or more pre-defined interventions will be included; |  |
| Comparator | Standard of care, no treatment or placebo; |  |
| Outcomes | • Mortality from all causes;<br>• Morbidity including : pulmonary, cardiac and renal complication rates, surgical complication rates (including readmissions);<br>• Patient reported experiences and |  |
|  | outcomes (PROMs/PREMS): pain-related outcomes (e.g. pain score rating, pain management satisfaction), quality of care (readiness for surgery, quality of care patient scores, quality of recovery after surgery);<br>Health service-related outcomes: length of stay (in hospital, in ICU) and economic/financial outcome; |  |
| Study design | Systematic reviews, meta-analysis<br>Randomized controlled trials<br>Non-randomized studies<br>Observational studies (cohort studies, case-control studies, cross-sectional studies, case series); | Case reports; |
| Study setting | Inpatient care (including patients whose condition requires admission to a hospital same day discharge surgical); | Outpatient clinics, medical and non-surgical management of spinal conditions; |
| Timing | Perioperative process- preadmission, preoperative, intraoperative and postoperative setting; | Studies incorporating long-term (greater than three months) postoperative rehabilitation; |

Outcome definitions in the review are in line with reported enhanced recovery pathways (34) (12). We defined our primary outcomes in the following groups:

- Morbidity, including pulmonary, cardiac, and renal complication rates; surgical complication rates; and readmission rates.
- Mortality from all causes.
- Patient-reported experiences and outcomes (PREMs/PROMs), including pain-related outcomes (pain score rating and/or opioid consumption, pain management satisfaction), readiness for surgery, quality of care patient scores, and quality of recovery outcomes.
- Health service-related outcomes, including length of stay and reported economic/financial outcomes (e.g., costs of the length of stay).

### Information sources and literature searches

We searched the following electronic databases (from 1990 onwards): MEDLINE via Ovid SP; EMBASE via Ovid SP; and Cochrane Library (Cochrane Database of Systematic Reviews and CENTRAL). In addition, we searched the grey literature through the following specific search engines: Google Scholar, OpenGrey, and GreyNet (35–37). We initiated the original search for studies in January 2020 and updated it in May 2020. For the search strategy, we combined keyword(s) and subject headings for all literature types in the pre-determined databases (29). We handled study overlap by tracking the index primary studies. For some selected pre-defined pathway components, there was a paucity of identified studies as pertaining to spinal surgery. Under those circumstances, we sought to identify large studies, meta-analysis or societal recommendations of best practice.

### Data extraction, management, analysis and presentation

Data were extracted from each study including publication details, study characteristics, participant characteristics, type of spinal surgery, intervention and comparator characteristics, and outcomes. The results of the data search were presented in a PRISMA flow diagram indicating the number of studies retrieved, screened and excluded as per exclusion criteria *(see Figure 1)*. We have presented our findings according to each individual predetermined element of the multimodal enhanced recovery pathway as presented in *Table 1*. For randomized controlled trials, one author extracted the information on the methodological quality of studies including random sequence generation, allocation concealment, blinding of participants and personnel, blinding of outcome assessment, incomplete outcome data, selective outcome reporting, and other bias (38) (39). For non-randomized studies data were unavailable/not collected on random sequence generation and allocation concealment.

### Risk of Bias and Thematic Synthesis

Risk of bias in randomized controlled studies was assessed using the Cochrane Risk of Bias tool (39). We used the ROBINS-I (Risk of Bias in Non-randomized Studies of Interventions) tool to assess the risk of bias in non-randomized studies (40). We used the revised AMSTAR-2 tool to assess the risk of bias in systematic reviews (41). Quality of evidence was classified according to the Grading of Recommendations, Assessment, Development and Evaluation (GRADE) system into one of four categories: high, moderate, low, and very low (42). Evidence based on randomized controlled trials was considered as high quality unless confidence in the evidence was decreased due to study limitations, inconsistency of results, indirectness of evidence, imprecision, and reporting biases. Observational studies were considered low quality; however, they were graded higher if the treatment effect observed is very large or if there is evidence of a dose-response relationship (33, 43)

Endpoint of the GRADE evidence summary consists of Evidence Profile(EP) tables across individual pathway components (42). We performed a thematic synthesis and narrative analysis for each proposed component of the complex intervention pathway (27). Quantitative data synthesis was not attempted due to the inherent heterogeneity of the studies with regard to design, populations, procedures, methods, and outcomes. This method of evidence synthesis is in line with other published enhanced recovery reviews (1, 13, 15, 17, 18, 44, 45). We did not make recommendations on the utility of pathway components.

## Results

Our search strategy retrieved a total of 5423 studies excluding duplicates using 22 different searches for the each relevant ERSS item as outlined in *Table 1*. Where studies were not available pertaining to predefined pathway component of spinal surgery, databases and grey literature were reviewed as relating to societal recommendations and major pertinent studies for perioperative patient management. This methodology yielded 148 further studies for inclusion. Following the initial search, we included 664 studies in the final review. The results of our search have been presented in *Figure 1, PRISMA Diagram*. We have grouped the evidence base according to the component of the pathway.

**Figure 1.**
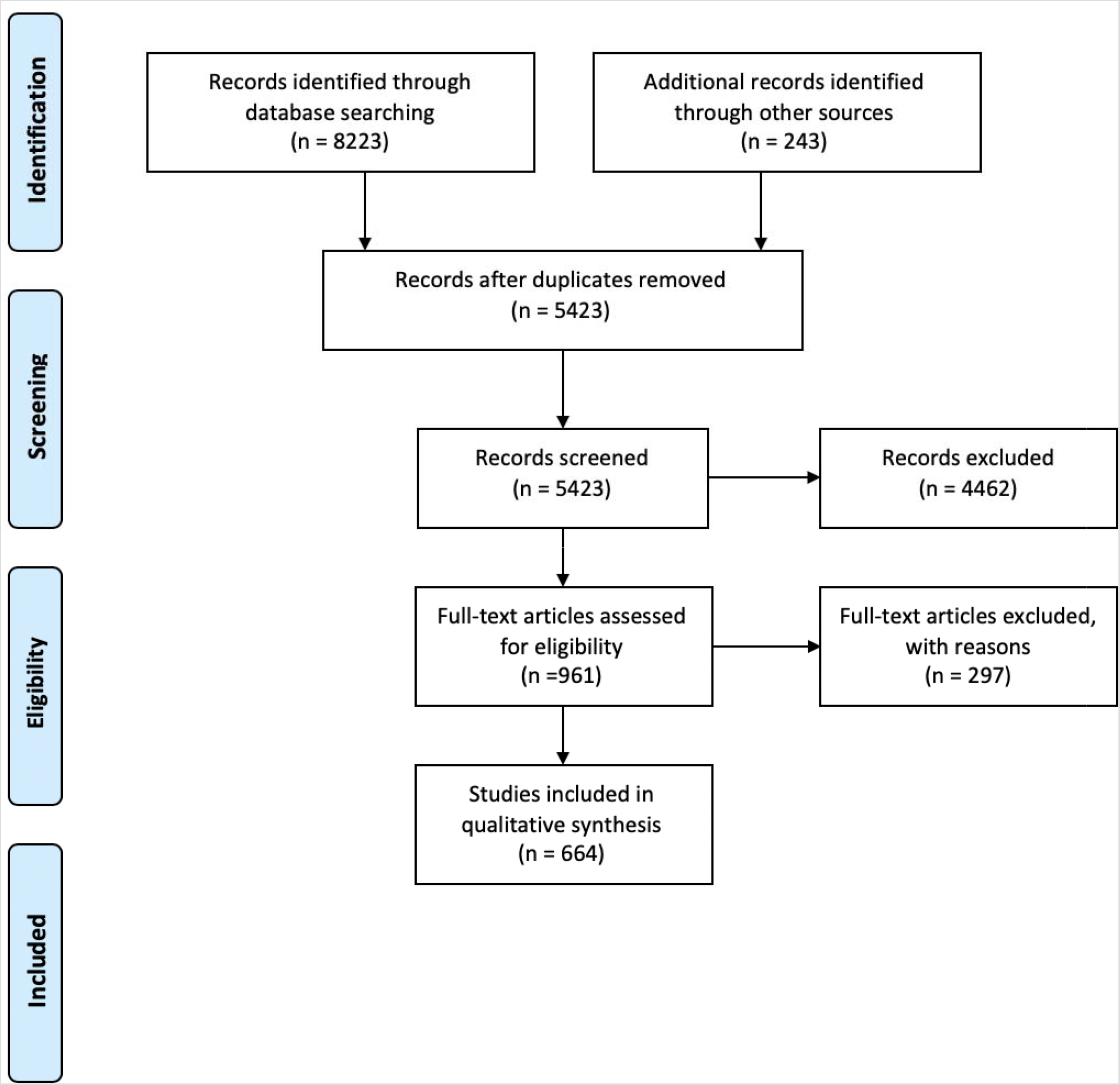
PRISMA diagram

Evidence Profile tables were generated when a number of studies were identified investigating an intervention for one of the predetermined outcomes. We have generated Evidence Profile tables for the following pathway components: 2.4 Tobacco use, 3.Prehabilitation, 4.1Preoperative nutritional screening, 5.Management of Anaemia, 6.Peri-operative blood conservation strategies, 8. Pre-emptive analgesia, 10.2 Antimicrobial prophylaxis, 11. Local anaesthetic infiltration, 12. Standard anaesthetic protocol, 13. Surgical access (open and minimally invasive spinal surgery), 16. Perioperative analgesia including use of intravenous lignocaine and 21.Mobilization.

For the following elements we identified published meta-analysis: 16. Perioperative analgesia including use of NSAIDS, ketamine, gabapentinoids and intrathecal morphine and 17. Thromboprophylaxis. We incorporated the relevant meta-analysis findings into each pathway.

We were able to identify heterogenous studies pertaining to surgery of the spine for the following components: 10.1 Surgical site preparation, 14. Maintenance of normothermia, 15. Intra-operative fluid and electrolyte therapy, 18. Urinary drainage, 19. Post-operative nutrition and fluid management and 20.Post-operative glycemic control. For these components, we performed a thematic synthesis.

Due to the paucity of evidence pertaining to spinal surgery, we identified societal recommendations for the following components: 1.Preadmission information, Risk assessment (2.1 Preoperative risk stratification, 2.2 Preoperative optimization and 2.3 Alcohol use), 4.2 Peri-operative immuno-nutrition, 6. Pre-operative fasting and carbohydrate loading, 9. Prevention of post-operative nausea and vomiting and 22. Audit.

### Presentation

We have presented our findings according to each individual element of the multimodal enhanced recovery pathway in line with other subspecialty ERAS pathways (13, 14, 46). We have grouped each element according to the chronological order of events with four distinct headings of preadmission, preoperative, intraoperative and postoperative elements.

## Discussion

### Preadmission period

The preadmission period is an opportunity for patient education, assessment of comorbidities, risk stratification and optimization of modifiable patient-related factors.

#### 1. Preadmission information, education and counseling

Patient information provision has long been considered a key element of enhanced recovery pathways (47). In a retrospective study, an in-person assessment at the pre-admission clinic was associated with a reduction in in-hospital mortality (48). Patient perioperative experience and the psychological aspect may be improved with pre-admission counseling (49–52). Psychopathology and patient expectations have been linked to poor results in spinal surgery with increased pain and decreased function. This has led to an increased reliance on pre-surgical psychological screening (PPS) as part of the surgical diagnostic process in spinal surgery (52–55).

Studies do not show any evidence of harm from preoperative information provision or psychological intervention. As such, there may be utility from information provision, balanced against no known harmful effects. There is limited evidence available pertaining to the intervention specifically in patients undergoing surgery of the spine.

#### 2. Risk assessment, preoperative assessment, optimization and lifestyle factor modification

##### 2.1 Pre-operative risk stratification

Perioperative period offers an opportunity to identify and manage fixed and modifiable patient risk factors (56). Nearly eighty percent of patient deaths come from the high-risk patient group (57). In a major retrospective study in the USA, it was found that if patients had a major complication within 30 days of surgery, the median survival was reduced by 69% at eight years (58). Multiple diverse risk scoring systems are currently in use for major surgery, including spinal surgery. Assessing cardiovascular risk can be undertaken whilst utilizing multivariate risk indices as incorporated in the ACC/AHA guidelines on perioperative cardiovascular evaluation and care for non-cardiac surgery (59). Lee’s cardiac risk index was designed and prospectively validated in patients undergoing non-urgent major non-cardiac surgery (60). Studies suggest that the Lee index is the most reliable generic perioperative cardiac risk index (61). Although Revised Cardiac Risk Index (RCRI) discriminates moderately well between patients with varying risks for cardiac complications, it performs poorly at predicting postoperative mortality (62). Of the general risk indices, POSSUM (Physiological and Operative Severity Scoring for the enumeration of Mortality and morbidity) has greater specificity for morbidity than mortality (63). The Portsmouth POSSUM (P-POSSUM) score has greater validity for mortality in the perioperative period (64). The American College of Surgeons National Surgical Quality Improvement Program (ACS NSQIP) has been deemed to have high internal validity (65). However external validations of the NSQIP have been inconsistent (66). A retrospective cohort study of the NSQIP database consisting of patients who had undergone elective posterior lumbar fusion was undertaken. The variables associated with greater risk and extended length of stay included increasing age, morbid obesity, operative time, multilevel procedure and intraoperative transfusion (67). In a retrospective review of NSQIP database, patients undergoing elective spinal surgery had the expected risk factors for cardiovascular complications consistent with those demonstrated by the Revised Cardiac Risk Index (68). Functional capacity is an important but subjective physician administered preoperative risk assessment tool (62). Duke Activity Status Index (DASI) scores were associated with predicting death or myocardial infarction within 30 days of surgery (69).

Individual preoperative risk assessment tools tests can be used to identify patients at risk of complications and perform a risk-based stratification. There is moderate quality evidence in support individualized risk stratification utilizing the most suitable risk-stratification assessment tool;

##### 2.2 Preoperative assessment and optimization

Patient preoperative assessment allows for an opportunity for examination of comorbidities with subsequent identification of fixed and modifiable comorbidities (70). Modifiable comorbidities may be amenable to improved perioperative management. Chronic conditions such as asthma, chronic obstructive airways disease, diabetes mellitus, anaemia and malnutrition should be optimized prior to surgery (71, 72). Obese patients having spinal surgery were found to have increased blood loss, prolonged hospital stay and were more likely to develop infection (73) (74). Patients with diabetes were found to have greater disability as compared to patients without diabetes in a 2-year longitudinal study of outcomes following spinal surgery (75). One year after undergoing spinal fusion, diabetic patients are more likely to have failed the procedure as compared to patients without diabetes (76) (77). Frailty is an emerging risk assessment tool, however definitions of this clinical entity are heterogenous (78). The degree to which preoperative optimization and modification of multimorbidity, affects healthcare outcomes is unclear (56, 78). Modifiable co-morbidities should be optimized using the preoperative process. Evidence base is of low quality due to a limited number of heterogenous studies.

##### 2.3 Alcohol use

Postoperative morbidity is increased by two- to threefold in alcohol abusers (79). Preoperative alcohol consumption is associated with an increased risk of postoperative morbidity, general infections, wound complications, pulmonary complications, prolonged stay at the hospital, and admission to intensive care unit (80). In a subset of patients without clinical or historical evidence of alcohol-related illness, one month of preoperative abstinence has been shown to significantly improve outcome (81) (80, 82). Significant alcohol consumption has been shown to be associated with increased perioperative morbidity. For alcohol abusers, 1 month of abstinence before surgery is beneficial. Evidence is considered to be of moderate quality due to heterogenous endpoints in available studies.

##### 2.4 Tobacco use

Smoking is an independent risk factor for non-union in spinal fusion procedures (83) (84) (85) (86). Post-operative infection and wound complications are significantly increased by tobacco consumption (85). Decreased risk of infection, perioperative respiratory problems, and wound complications have been demonstrated one month after cessation of smoking(87). Longer periods of cessation of smoking appear to be more effective in reducing the incidence/risk of postoperative complications (88, 89). There is translational high quality evidence for cessation of smoking at least 4 weeks pre-operatively.

#### 3. Prehabilitation

Prehabilitation can be defined as “the process of enhancing the functional capability of an individual in preparation for the surgical intervention”. This process consists of: functional preoperative prehabilitation, nutritional and psychological intervention (90). Pre-operative functional capacity is closely related to post-operative morbidity (69). Whether improving the post-operative outcomes through prehabilitation has beneficial effects on mortality is not yet clear (91). A randomized clinical trial of multimodal prehabilitation in spinal surgery found that the intervention group reached the recovery milestones earlier and left the hospital earlier (92). A cohort study with a focus on functional prehabilitation identified an appreciable improvement in patient satisfaction scores in the study group (93). Health-economic benefits have been greater in patients having prehabilitation (94) (93). Patient reported outcomes such as readiness for surgery and perceived quality of life, were found to be improved by pre-operative neuroscience education and physiotherapy (95) (96, 97). Overall evidence quality for prehabilitation is moderate.

#### 4. Preoperative Nutritional Care

##### 4.1. Nutritional Assessment and Screening

Preoperative malnutrition as defined by hypoalbuminaemia, has been shown to be an independent risk factor for increased postoperative complication rates, including cardiorespiratory problems, increased rate of infection and unplanned readmission within 30 days post discharge after elective spinal surgery (98) (98) (99, 100) (101). Well-known risk factors for nutritional depletion in spinal surgery include: diagnosis of cerebral palsy, circumferential spinal surgery, fusion levels greater than or equal to 10, and age over 50 (85) (102).

There is moderate quality evidence available for performance of risk assessment and screening of nutritional status in patients undergoing spinal surgery.

##### 4.2. Perioperative immuno-nutrition

Overall systematic evidence on immuno-nutrition (IN) in surgery has been contradictory (103) (104). Clinical studies demonstrating benefit of IN are heterogenous with non-standardized primary solutions, controls or timing of administration of supplements (105). There is no evidence for use of IN in patients undergoing surgery of the spine.

#### 5. Management of anaemia

Preoperative anemia is an independent risk factor for increased 30-day mortality and morbidity in surgical patients (106) (107) (108, 109). In patients undergoing surgery of the spine, preoperative anaemia was associated with increased length of stay(110, 111). Intraoperative blood transfusion in spinal surgery has been associated with increased postoperative complications, length of hospital stay and 30-day re-admission rates (112). It is however unclear whether correcting iron deficiency anaemia improves reported outcomes, other than decreasing the risks associated with perioperative blood transfusion (113) (114). It is unknown whether correcting non-anaemic iron deficiency (NAID) decreases the risk of perioperative complications (115). The association of iron replacement therapy, in particular intravenous format, with infection, is currently contentious (116) (117).

Clinically guided appropriate pre-operative use of intravenous or oral iron, vitamin B12, folic acid or erythropoietin for patients suffering from anaemia and/or low iron stores should be implemented in patients undergoing moderate and major spinal surgery. There is moderate quality translational evidence for correcting the iron deficiency anaemia, in order to decrease the perioperative risk of complications secondary to blood transfusion.

#### 6. Perioperative blood conservation strategies

Patients undergoing moderate and major spinal surgery are at risk of significant blood loss necessitating fluid and blood product replacement (118). With attenuating blood loss, risks of attendant transfusion can be minimized.

Blood loss management in spinal surgery can be approached through three pillars of patient blood management

1. detection and treatment of preoperative anaemia (see above).
2. reduction of perioperative red blood cell loss.
3. harnessing and optimizing the patient-specific physiological reserve of anaemia (including restrictive transfusion triggers) (119).

Recent systematic analysis of tranexamic acid use in spinal surgery patients concluded that the use of tranexamic acid (TXA) was effective in reducing intra-operative blood loss and decreased the volume of blood transfusion (120). It is likely that the higher bolus doses employed (greater or equal to 15mg/kg followed by intraoperative infusion) were more effective in attenuating blood loss and transfusion requirements (121, 122). Lower perioperative blood loss has consistently been demonstrated with the use of £-amin-caproic acid (EACA) in spinal surgery (121). Point of care testing devices allow for standardization of transfusion practices and early identification and treatment of hypofibrinogenemia (119). In a study of patients undergoing major spinal surgery ROTEM (ROtational ThromboElastoMetry) device used with TXA was found to lead to a significantly lower blood loss and lower transfusion of packed red blood cells as compared to the TXA alone (123). Other studies have demonstrated the utility of ROTEM in decreasing the rate of blood product transfusion (124, 125).

Blood conservation options include preoperative autologous blood donation and intraoperative cell saver use.

Preoperative autologous blood donation in elective major spine surgery has been effective in reducing allogenic transfusion, however inclusion in the program resulted in increased risk of transfusion (126) (127) (128). A Cochrane meta-analysis assessing the use of cell saver in major surgery including heterogenous orthopaedic surgery, demonstrated statistically significant decreased rate of allogenic blood transfusion (129). The use of the cell saver in posterior spinal instrumentation and fusion surgery in school-aged children and adolescents was able to decrease the amount of intraoperative allogeneic RBC transfusion but failed to decrease total perioperative allogeneic RBC transfusion (130) (131). A single retrospective case series found cell-saver use to be associated with increased risk of bleeding in patients having spinal fusion surgery (132). There is no evidence supporting the use of controlled hypotension to minimize the risk of bleeding, particularly in prone patients. By contrast, in a large retrospective cohort study of all patient populations undergoing a range of surgical procedures, significant hypotension was found to be associated with increased 30-day mortality (133). Perioperative blood conservation is aided through simple clinical measures such as temperature regulation, optimal patient positioning and meticulous surgical techniques.

There is high quality evidence for antifibrinolytic use in surgery of the spine when significant blood loss is anticipated. There is moderate quality evidence for cell saver use when significant blood loss is anticipated. There is low quality evidence for use of point of care testing to decrease the number of red blood cell units transfused.

### Pre-operative period

#### 7. Preoperative fasting and carbohydrate loading

In a randomized controlled trial examining patient population having coronary artery bypass grafting or spinal surgery, preoperative carbohydrate loading did not attenuate postoperative insulin sensitivity (134). This is contradictory to other general surgical trials which demonstrate improved insulin postoperative sensitivity with CHO loading (135) (136). Drinking carbohydrate-rich fluids before elective surgery improves subjective well-being, reduces thirst and hunger and reduces postoperative insulin resistance but has no significant effect on length of postoperative stay and mortality (137, 138). The clinical relevance of administering preoperative CHO loading in patients with diabetes remains to be established (45). Permitting patients to drink water or clear fluid preoperatively results in significantly lower gastric volumes (136). International guidelines allow for unrestricted intake of clear fluids up to two hours before elective surgery in patients not considered to have impaired gastric emptying (137) (139).

In spinal surgical patients without delayed gastric emptying standard societal fasting implementations can be made. Patients should be allowed to eat up until 6 h and take clear fluids including CHO drinks, up until 2 h before initiation of anaesthesia. Preoperative treatment with oral CHOs may not be suitable in patients with documented delayed gastric emptying, gastrointestinal motility disorders and in patients undergoing emergency surgery.

#### 8. Pre-emptive analgesia

A number of studies found that pre-emptive administration of gabapentin reduced the opioid consumption and pain scores in the postoperative period in spinal patients (140) (141–143). The most effective dose in lowering the post-operative pain scores was found to be 600 mg (141). Impact of multimodal anti-inflammatory regimes combined with gabapentinoids, is significant in lowering the postoperative pain scores (144) (145). Parecoxib and ketorolac were found to be equally effective in improving postoperative pain measures. Both were superior to placebo in patients undergoing posterior lumbar fusion (146) (146). Pre-emptive epidural analgesia for thoracolumbar spine surgery has not been deemed effective (147).

Multimodal pre-emptive analgesia utilizing individual gabapentinoids and/or non-steroidal antiinflammatory agents improves pain scores and functional measures in the immediate post-operative period. There is high quality evidence for preemptive administration of gabapentinoids in patients undergoing surgery of the spine.

There is moderate quality evidence for pre-emptive administration of multimodal anti-inflammatory regimes combined with gabapentinoids. Evidence quality is low for sole administration of individual antiinflammatory agents.

### Intraoperative period

#### 9. Prevention of nausea and vomiting (PONV)

Postoperative nausea and vomiting (PONV) are amongst the most frequent postoperative complications, impacting the quality of recovery and causing patient dissatisfaction (148). Patients should be risk-stratified according to the baseline risk of PONV (149). Female, non-smokers and patients with a history of motion sickness are at greatest risk of PONV. Anaesthesia related factors increasing the risk of PONV include the use of volatile anaesthesia, nitrous oxide and opioid administration. Longer surgical times are associated with greater risk of PONV. In patients undergoing spinal surgery, particular risk factor for increased PONV is the need for significant intraoperative and postoperative opioid administration. Standard societal guidelines for PONV prophylaxis and management apply to patients undergoing spinal surgery (148). With non-pharmacological measures, avoidance of fasting and dehydration has been recommended. Multi-modal analgesia with opioid sparing effect has a beneficial influence on the risk of PONV (150) (151). With regards to pharmacological management, single agent prophylaxis is adequate in most patients. In patients who are at risk of moderate to severe PONV, dual agent PONV prophylaxis is recommended. In patients with three or more risk factors, the recommended approach is to administer total intravenous anaesthesia.

Risk assessment of patients according to the anaesthetic and procedural factors is recommended. Stepwise non-pharmacological and pharmacological PONV prophylaxis according to the guidelines is recommended. Use of anaesthetic techniques which minimize risk of PONV in high-risk patients should be considered.

There is high quality evidence for risk stratification of patients and appropriate anti-emetic prophylaxis. There is moderate quality evidence for opioid sparing techniques as well as avoidance of nitrous oxide and volatile anaesthesia;

#### 10. Surgical site preparation and antimicrobial prophylaxis

##### 10.1. Surgical site preparation

Reported post-operative infection rates in spinal surgery range from two to thirteen percent (152). Large prospective cohort studies have demonstrated that the surgical site infection rate is equivalent with both topical chlorhexidine gluconate and povidone in spinal surgical patients (153) (154). Topical chlorhexidine with alcohol compared to povidone alone, decreased the bacterial load significantly in spinal surgery (155) (156). A review evaluating randomized controlled trials in all types of surgeries concluded that alcohol-based agents are superior to aqueous solutions (157).

There is high quality evidence for using alcohol-based preparations. There is moderate quality evidence for decreasing the viable bacterial load utilizing CHG with alcohol solution.

##### 10.2 Antimicrobial prophylaxis

Risk factors for surgical site infections after spinal surgery include extended duration of procedure (longer than two hours), excessive blood loss (greater than one liter), staged procedure, multilevel fusion, foreign body placement, combined, anterior and posterior fusion, and poor peri-operative glycemic control (158). Surgical site infection is less likely in procedures at the cervical spine level or with an anterior surgical approach (159). Current guidelines recommend intravenous cephazolin as the first choice agent for antimicrobial prophylaxis for most surgical procedures (160). In patients with MRSA, intravenous vancomycin is recommended one hour prior to skin incision. Clindamycin is an acceptable alternative in patients with a cephalosporin or vancomycin allergy. In the setting of risk for SSI due to gram-negative pathogens, an additional agent may be warranted (such as an aminoglycoside, aztreonam, or a fluoroquinolone). In order to ensure adequate antimicrobial serum and tissue concentrations, repeat intraoperative dosing is warranted for procedures that exceed two half-lives of the drug and for procedures in which there is excessive blood loss (160). In a meta-analysis incorporating 6 prospective randomized-controlled trials, antibiotic prophylaxis was found to decrease the rate of infection (161). Whether postoperative infections are reduced by continuing use of prophylactic antibiotics remains controversial (162). In a meta-analysis consisting of 14 mostly class 3 evidence studies, vancomycin powder was found to decrease the likelihood of surgical site infection (163). Vancomycin powder was found to decrease the rate of deep space infections requiring re-operation (164). Vancomycin powder should be restricted to procedures and patients most at risk of MRSA-related surgical site infection (165) (166) (167).

Routine prophylaxis with cefazolin within 1 hour prior to skin incision is recommended. Patients with MRSA should be treated prophylactically with vancomycin initiated 1 hour prior to skin incision. There is high quality evidence for intra-operative antibiotic prophylaxis. There is low quality evidence for use of intravenous vancomycin in patients at risk of MRSA.

#### 11. Local anaesthetic infiltration

The benefits of intra-operative wound infiltration for postoperative analgesia in spinal surgery are controversial. A number of studies have demonstrated conflicting results in this area (168–171). A metaanalysis of nine trials exploring the effect of wound infiltration in spinal surgery concluded that only a few trials observed a mild to modest pain score reduction. Of the trials which did show pain reduction, the analgesic benefit was noted in the immediate post-operative period (172). Local anaesthetic wound infiltration in major spinal surgery has some immediate benefit on postoperative pain scores. There is moderate quality evidence for intra-operative administration of long-acting local anaesthetic administration.

#### 12. Standard Anaesthetic Protocol

Prior systematic reviews and meta-analysis have concluded that recovery parameters are improved with the use of total intravenous anaesthesia (TIVA)(173) (174). There is some evidence that patients receiving TIVA had improved cognitive outcomes in post-anaesthesia recovery unit in all types of surgical patients (175). Patients anesthetized with propofol-based TIVA reported less pain during coughing and consumed less daily and total PCA fentanyl after lumbar spine surgery (176). This finding was not consistent across all studies (177). Remifentanil, ultra-short acting phenyl-piperidine derivative is used in spinal surgery as part of total intravenous anaesthesia or inhalational anaesthesia protocols. Indications for use in spinal surgery include: improved endotracheal tube tolerance, improved surgical conditions and facilitations of peripheral neuromuscular monitoring. Severe postoperative pain after the intraoperative use of remifentanil has repeatedly been linked to the development of acute tolerance and/or opioid induced hyperalgesia (178). In patients undergoing spinal fusion remifentanil dosage up to 0.16 mg/kg/min did not cause an increased post-operative opioid consumption (179). In contrast in patients having correction of scoliosis where higher doses of remifentanil of 0.28 mcg/kg/min were used for longer duration, the requirements for post-operative analgesia were thirty percent higher in the remifentanil group(180). Neurologic monitoring in spinal surgery is performed using the intraoperative somatosensory potentials (SSEP’s) and/or the Motor Evoked Potentials (MEP’s). During the SSEP monitoring anaesthetic drugs produce a dose dependent increase in latency and a decrease in amplitude. The overall quality of SSEP is superior when propofol total intravenous anaesthesia is used. International Society of Intraoperative Neurophysiology recommends use of propofol and opioid (181). MEP’s display extreme sensitivity to the inhibitory effects of volatile agents even at concentrations as low as 0.25 MAC. Due to a lower level of interference with monitoring MEP’s, propofol total intravenous anaesthesia is recommended for patients requiring spinal cord neurophysiological monitoring during surgery. There is moderate quality evidence for use of total intravenous anaesthesia in patients undergoing surgery of the spine. There is low quality evidence for continuous intra-operative remifentanil infusion use in spine surgery.

#### 13. Surgical access (open and minimally invasive spinal surgery, including robotic surgery)

Minimally Invasive Spinal Surgical (MISS) techniques can be viewed as a critical component of enhanced recovery in spinal surgery protocols (ERSS) (182). MISS techniques have been efficacious in decreasing postoperative pain in observational studies (183). Wang et all used minimally invasive surgical approaches in awake patients within the specific context of an ERSS protocol (184). With a focus on minimally invasive transcutaneous lumbar inter-body fusion, the authors showed that ERAS in this group of patients was feasible and afforded improved early functional outcomes. MISS approach studied within the enhanced recovery protocol was found to be effective in oncological spinal patients, where it was found to decrease the pain scores and lower the opioid consumption (185). Faster mobilization by a factor of two was demonstrated in patients undergoing minimally invasive thoracic inter-lumbar body fusion compared to open procedure (186). Reduced length of stay together with significant cost saving has been identified in studies utilizing MISS techniques (32). In contrast to single studies and qualitative reviews, a quantitative meta-analysis found there was equipoise in patients undergoing lumbar minimally invasive procedures (187). A multicenter study found equivalent outcomes for obese patients having spinal MISS or open techniques (188). Conversely, Senkar et all found minimally invasive surgical techniques had the highest utility in patients with multiple comorbidities (189). There is evidence that minimally invasive surgical approaches improve pain scores, decrease opioid consumption and decrease length of stay, when used within the appropriate clinical context. There is moderate quality evidence for the intervention in appropriate clinical context.

#### 14. Maintenance of Normothermia

Maintenance of normothermia has been shown to decrease the frequency of morbid cardiac events and the rate of blood product transfusion in major surgery (190, 191). In spinal procedures with potential neurological cord compromise, maintenance of normothermia and avoidance of hyperthermia is recommended (192). There is little scientific literature supporting the neuroprotective effects of hypothermia on the spinal cord in elective or emergency spinal surgery (193). In pediatric spinal surgery maintenance of normothermia was found to be associated with a lower allogenic red blood cell transfusion rate (194). In a retrospective study intraoperative hypothermia was associated with a 30 percent lower rate of acute kidney injury in spinal surgery under general anaesthesia (195). Measures to maintain normothermia should be implemented in spinal surgical patients. There is moderate quality evidence for maintenance of intraoperative normothermia.

#### 15. Intraoperative fluid and electrolyte therapy

For the minor range of spinal surgeries intraoperative fluid management goals are achievable with routine monitoring. In major surgery, goal-directed therapy has been recommended (196). The use of advanced haemodynamic monitoring equipment may enhance clinical decision-making when used in comparison to conventional monitoring (196). Advanced haemodynamic monitoring equipment chosen should be based on a clinical risk-management strategy and patient, anaesthetic, surgical and institutional factors. A prior meta-analysis has demonstrated that the use of pre-emptive hemodynamic monitoring and proactive therapy reduced both the mortality and complication rates in major surgical procedures (197). In a large heterogenous meta-analysis goal-directed fluid management decreased the incidence of complications and duration of stay (198). In a retrospective observational trial in patients undergoing prone spinal surgery, goal directed fluid management was found to decrease blood loss and transfusion, improve postoperative respiratory performance and allow for faster return of bowel function (199). In a retrospective cohort study in spinal patients the liberal fluid strategy was associated with an increased rate of pulmonary complications (200). It is currently unclear whether crystalloids or colloids should be used as a primary maintenance and replacement fluid in spinal surgery.

Goal-directed fluid management may decrease the rate of complications and duration of stay when implemented in the appropriate clinical context. There is low quality evidence for goal-directed intraoperative fluid management using contextually appropriate indicators and measurements of cardiac output in patients undergoing major surgery of the spine.

#### 16. Peri-operative analgesia

Poorly controlled pain in the post-operative period can influence mobility and result in increased rate of complications of deep venous thrombosis, pulmonary embolism and pneumonia (201).

### NSAIDS and acetaminophen

A recent meta-analysis of eight trials identified that NSAIDs are effective in postoperative analgesia after lumbar spine surgery. The study found that NSAID dose, different surgery types, and analgesic type might influence the efficacy of NSAIDs (202). A meta-analysis of 17 studies demonstrated that addition of NSAIDs to opioid analgesics alone resulted in lower pain scores and less morphine equivalents consumed (203). In a meta-analysis of seven spine fusion studies, no statistically significant association between NSAID exposure and nonunion was identified (odds ratio = 2.2, 95% confidence interval 0.8–6.3) (204). It is likely that adverse effects of NSAID’s on bone healing/fusion in adult spine surgery are dose-dependent (205). While there is limited evidence for the use of acetaminophen specifically in spinal surgery, it is a well-established analgesic agent for a wide range of related surgeries (201).

### N-methyl D-aspartate antagonists

Randomized controlled trials demonstrating decreased opioid consumption and lower pain scores following intraoperative and post-operative ketamine (206, 207) (208). These findings are in line with a meta-analysis of eight trials (209). A single study showed no benefit of low dose ketamine in major lumbar surgery (210).Methadone and magnesium through their NMDA antagonism may also be of benefit; however, data are limited and further studies are indicated (211) (212, 213).

### Alpha-2 receptor agonists

Data supporting the use of alpha-2 receptor agonists in major spine surgery are limited; studies have demonstrated conflicting findings (214) (215).

### Gabapentinoids

In a systematic review and meta-analysis by Yu et al, perioperative administration of gabapentinoids was found to decrease opioid consumption and pain intensity in the immediate post-operative period (142). Other high quality prospective studies have deemed gabapentinoids effective at reducing the opioid consumption when continued for at least 24 hours post-operatively (143). A prospective, double-blind study, randomized control trial by Khurana et al. showed a stronger benefit for pregabalin over gabapentin versus placebo for pain and functional status in the post-operative period and at 3 months (216).

### Intravenous lignocaine

In a number of controlled trials in both adult and pediatric major and minor spine surgery, perioperative lignocaine infusion was demonstrated to improve pain scores and decrease opioid consumption (217–220). Conversely, in a randomized controlled trial of seventy patients undergoing posterior spine surgery, there was no analgesic benefit of a systemic lignocaine infusion as compared to placebo (221).

### Regional analgesia

Intrathecal morphine administration in a wide dosage range as a single injection has been found to be effective as a postoperative analgesic in spinal surgery, though doses greater than six mcg,kg^−1^ are associated with postoperative respiratory depression (222, 223) (224–229). In a meta-analysis of eight randomized controlled trials, intrathecal morphine was an effective analgesic (230).

### Multimodal regimens

Prior review articles have highlighted multimodal analgesia as a key to enhanced recovery in spinal surgery (201) (231) (5). Multimodal analgesia bundles have been incorporated into most care pathways of enhanced recovery in spinal surgery (9, 12, 232). A numbed of retrospective studies have demonstrated decreased pain measurement outcomes including post-operative opioid consumption (233) (234–236). In contrast to other studies, a single randomized controlled trial of optimally dosed multimodal regime did not show any benefit over the placebo components when evaluated in terms of quality of recovery scores or analgesic components (237). Minimally invasive opioid free enhanced recovery protocols in spinal surgery have been shown to have a favorable profile on perioperative opioid consumption (238).

Simple analgesics such as acetaminophen and NSAIDs are safe and efficacious, particularly in combination. There is high quality evidence for perioperative administration of NSAID’s. Ketamine in both intraoperative and post-operative infusions, reduces pain scores, opioid requirements in the immediate and late post-operative phases. There is moderate quality for intraoperative ketamine administration. There is very low quality evidence for administration of other NMDA antagonists and alpha-2-agonists. There is high quality evidence for perioperative gabapentinoid administration. Consideration should be given to perioperative intravenous lignocaine infusion administration. There is moderate quality evidence for perioperative intravenous lignocaine administration. There is moderate quality evidence for use of intrathecal morphine in spinal surgery, although its utility may be limited by logistical factors. Clinically appropriate multimodal opioid-sparing regimens should be considered in all patients undergoing spine surgery. There is moderate quality evidence for instituting peri-operative multimodal analgesia.

### Postoperative period

#### 17. Thromboprophylaxis

Mechanical thromboprophylaxis is a proven measure to decrease the risk of deep venous thrombosis (DVT) in the absence of chemoprophylaxis (239). A meta-analysis conducted in 2018 found that the incidence of DVT and pulmonary embolism (PE) in spinal surgical population was relatively low regardless of prophylaxis type. The authors commented that there was a higher mean incidence of DVT and PE in the mechanoprophylaxis group (DVT: 1%, PE: 0.81%) compared to the chemoprophylaxis group (DVT: 0.85%, PE: 0.58%) (240). In this study, when PE occurred it was fatal in six percent of patients. Perception of true incidence of post-operative epidural haematoma in spinal surgical patients is varied (241).

Patients undergoing spinal surgery should have mechanical thromboprophylaxis by well-fitting compression stockings and/or intermittent pneumatic compression until discharge. There is moderate quality evidence for postoperative mechanical thromboprophylaxis in patients undergoing spinal surgery. There is low quality evidence for postoperative chemical thromboprophylaxis in patients undergoing spinal surgery.

#### 18. Urinary drainage

Urinary catheter use beyond 48 hours following surgery has been associated with an increase in hospital-acquired urinary tract infections and 30-day mortality (242).In (242).In a nested cohort study in a neurological intensive care unit, an increased rate of urinary infection was noted in patients, where catheter remained in place for longer than seven days (243). Risk factors for postoperative urinary retention in spinal surgery include older age, benign prostatic hypertrophy, chronic constipation, longer duration of surgery and posterior spinal fusion (244) (245) (246).

If urinary drainage is indicated, the duration of catheterization should be individualized based on known risk factors for urinary retention. There is moderate quality evidence for urinary catheter removal within 48 hours after surgery.

#### 19. Postoperative nutrition and fluid management

Many of the studies report on early mobilization in conjunction with dietary libertization (183, 247). When performed together, the two can reduce length of stay and costs without increasing early or late complications in adolescents undergoing posterior spinal fusion (248). Results of the RELIEF trial suggest that we should be more cautious with postoperative restrictive fluid strategies in patients having major abdominal surgery (249). In patients having major spine surgery, goal orientated post-operative fluid management may be more appropriate than a restrictive approach, although specific evidence is currently lacking. Intraoperative haemodynamic framework utilized intra-operatively may be continued into the post-operative period in the high-risk patient group or those with intraoperative complications. In line with other ERAS guidelines patients should be encouraged to transition as early as tolerated to oral intake.

#### 20. Postoperative glycemic control

A retrospective cohort study incorporating population undergoing spine surgery found that perioperative hyperglycemia increases the risk of adverse post-operative events in the non-diabetic patient group (250). Tighter glycemic control may mitigate the risk of surgical site infection in patients with diabetes (251). There remains insufficient evidence that strict glycemic control is advantageous over conventional management for prevention of surgical site infection (252). Although it is clear that perioperative hyperglycemia is deleterious, the optimal management paradigm in the postoperative period remains uncertain (251).

Severe perioperative hyperglycemia is deleterious, however optimal blood glucose management paradigm in the postoperative period remains uncertain. It is prudent to maintain more conventional blood glucose target in the postoperative period in patients undergoing spinal surgery. There is low quality evidence for conventional postoperative blood glucose control.

#### 21. Early Mobilization

Early mobilization is thought to be a key component of ERSS (12) (253). There is no clear definition of mobilizing, which may include simple exercise in bed, walking in the room or walking further distances (45). The overall outcome of these pathways has been that of significant decreased length of stay; as well as improved patient satisfaction measures in selected studies (183) (254) (232, 247, 255, 256). A study focusing on behavioral outcomes of early mobilization and rehabilitation education, identified decreased postoperative patient anxiety and enhanced self-care ability (257). A narrative review explored the impact of structured goal-directed mobilization on recovery parameters following spinal surgery. Reduced complication rates, improved patient-reported outcomes and decreased length of stay were noted in the patients undergoing early mobilization(258).

Patients should be encouraged to mobilize actively on the day of surgery as guided by clinical condition and surgical concerns. In the absence of a clear definition of early mobilization, institutions should be encouraged to set their own benchmarks. There is moderate quality evidence for the intervention due to the imprecision in defining mobilization as well as retrospective nature of studies.

### Quality of care measures

#### 22. Audit

Systematic audit is the preferred practice pattern in order to review compliance rates with the ERAS implemented interventions (259). There is evidence in retrospective studies that greater compliance with ERAS processes and protocols improves desired perioperative outcomes (260–263). There is a paucity of audit data in multimodal ERSS protocols.

Audit performance of ERAS as a part of a quality improvement cycle increases the compliance with the multi-disciplinary program and results in improved measured outcomes.

**Table 3.** Summary of the recommendation for clinical care

| *ERSS Element* | *Recommendation for clinical care* |
| --- | --- |
| 1. Preadmission information, education and counseling | Preadmission information, education and counseling may have a positive impact on subjective perioperative patient experience. Studies do not show any evidence of harm; |
| 2. Risk assessment, preoperative optimization, including lifestyle factor modification |  |
| 2.1 Preoperative risk stratification | Preoperative risk assessment tools tests can be used to identify patients at risk of complications; Prognostic accuracy and predictive ability of a risk measurement tool should be considered; |
| 2.2 Preoperative assessment and optimization | Preoperative assessment and optimization of modifiable comorbidities should be performed on all patients. Although the degree to which preoperative optimization affects healthcare outcomes is unclear, it is intuitive that any modifiable co-morbidities should be optimized using the preoperative process; |
| 2.3 Alcohol use | Increased alcohol consumption has been shown to be associated with increased perioperative morbidity. For alcohol abusers, 1 month of abstinence before surgery is beneficial; |
| 2.4 Tobacco use | For tobacco users, 1 month of abstinence before surgery decreases the risk of infection and wound healing; |
| 3. Prehabilitation | Multimodal prehabilitation may improve patient reported outcome measures and allow for earlier hospital discharge in spinal surgery; |
| 4. Preoperative Nutritional Care |  |
| 4.1. Nutritional Assessment and Screening | Risk assessment and screening of nutritional status should be performed in patients undergoing spinal surgery; |
| 4.2 Immuno-nutrition | There have been no benefits of immuno-nutrition in spinal surgery; |
| 5. Management of anaemia | Clinically guided use of intravenous or oral iron, vitamin B12, folic acid or erythropoietin for patients suffering from anaemia and/or low iron stores should be implemented in patients undergoing moderate and major spinal surgery; |
| 6. Perioperative blood conservation strategies | Tranexamic acid used at the higher dosage is effective in decreasing intraoperative blood loss. Cell saver techniques should be used in adolescents undergoing major corrective surgery. Cell-saver techniques may be beneficial when major blood loss is anticipated in adults. |
| 7. Preoperative fasting and carbohydrate loading | In patients without delayed gastric emptying standard societal fasting implementations can be made; |
| 8. Pre-emptive analgesia | Multimodal pre-emptive analgesia utilizing individual gabapentinoids and non-steroidal anti-inflammatory agents improves pain scores and functional measures in the immediate post-operative period; |
| 9. Prevention of postoperative nausea and vomiting | Risk assessment of patients according to the anaesthetic and procedural factors is recommended. Step-wise non-pharmacological and pharmacological PONV prophylaxis according to the guidelines is recommended. Use of anaesthetic techniques which minimize risk of PONV in high-risk patients should be considered; |
| 10. Surgical site preparation and antimicrobial prophylaxis |  |
| 10.1 Surgical site preparation | Chlorhexidine gluconate (CHG) is more effective at reducing the pre-operative viable bacterial load than povidone. Alcohol based agents are superior to aqueous solutions; |
| 10.2 Antimicrobial prophylaxis | Routine prophylaxis with cefazolin within 1 hour prior to skin incision is recommended. Patients with MRSA should be treated prophylactically with vancomycin initiated 1 hour prior to skin incision; |
| 11. Local anaesthetic infiltration | Local anaesthetic wound infiltration in major spinal surgery has some immediate benefit on postoperative pain scores; |
| 12. Standard Anaesthetic protocol | Total intravenous anaesthesia utilizing propofol demonstrates improved post-operative recovery markers after surgery. Higher total doses of intra-operative remifentanyl are likely to result in the phenomena of acute opioid tolerance and hyperalgesia; |
| 13. Surgical access- open and minimally invasive spinal surgery | Minimally invasive surgical approaches improve pain scores, decrease opioid consumption and decrease length of stay, when used within the appropriate clinical context; |
| 14. Maintain normothermia | Measures to maintain normothermia and avoid hyperthermia should be implemented in spinal surgical patients; |
| 15. Intraoperative fluid and electrolyte therapy | Goal-directed intraoperative fluid management should be implemented using contextually appropriate indicators and measurements of cardiac output in patients undergoing moderate/major surgery of the spine; |
| 16. Peri-operative analgesia | Simple analgesics such as acetaminophen and NSAIDs are safe and efficacious, particularly in combination. Ketamine in both intraoperative and post-operative form reduces pain scores. Consideration should be given to pre-emptive gabapentinoid |
|  | administration; Intravenous lignocaine has been shown to have immediate and long-term benefits for analgesia and function; |
| 17. Thromboprophylaxis | Patients undergoing spinal surgery should have mechanical thromboprophylaxis by well-fitting compression stockings and/or intermittent pneumatic compression until discharge. There is a role for careful use of chemoprophylaxis; |
| 18. Urinary drainage | Urinary catheters should be removed as soon as feasible; |
| 19. Post-operative nutrition and fluid management | Patients should be encouraged to transition as early as tolerated to oral intake. Postoperative fluid replacement should be carefully guided by patient intake and ongoing fluid losses; |
| 20. Post-operative glycemic control | Maintain conventional blood glucose target in the postoperative period in patients undergoing spinal surgery; |
| 21. Early mobilization | Patients should be mobilized actively on the day of surgery as permitted by the clinical condition; Patients should be encouraged to mobilize actively from the morning of the first postoperative day; |
| 22. Audit | Audit of compliance and care outcomes should be performed regularly in ERSS programs; |

### Implications of this Study and Future Directions

We have identified, delineated and presented the evidence base for first comprehensive multimodal program for Enhanced Recovery in Spinal Surgery (ERSS). A continuous issue when discussing enhanced recovery protocols is that of contention as to which components have the highest clinical utility, accompanied by somewhat arbitrary decisions on incorporating different elements into the program. We identified a high level of evidence for administration pre-emptive analgesia, peri-operative blood conservation (antifibrinolytic use), surgical site preparation and antibiotic prophylaxis.. Although evidence base for cessation of smoking in surgery of the spine is low, there is translational high level evidence from other surgical specialties. In contrast with prior ERSS reviews we identified moderate evidence base for utilization of minimally invasive surgery and use of multimodal analgesia (12). Although early mobilization and dietary libertization are considered critical in enhanced recovery, we identified a moderate level of evidence for institution of these interventions. This finding may be confounded by difficulties instituting high quality studies in these domains. Some clinical units may choose to use certain aspects of this proposed perioperative program as suited best to their unique location and practice pattern. Many of the studies illustrating institutional implementation of some components of ERSS have shown significant reduction in length of stay and consequent improved financial outcome (6). The most comprehensive ERSS program was instituted in patients having degenerative spinal surgery. Over the five-year period, the authors demonstrated a clear trend toward a higher proportion of patients discharged home after a one-night stay, with a concomitant decrease in adverse events in the overall cohort of 2592 patients (9).

Evidence base is low in certain research areas. Most of the studies assessed were conducted outside the context of enhanced recovery program. This may have a negative bias effect, where the effect of an individual component may be higher than estimated when used within the ERSS pathway. Their combination with other components in a particular pathway is thought to have a synergistic effect. In addition to including studies focusing on individual ERSS elements, we evaluated studies focusing on bundles of care. Through the additive incremental value of each component, this may have a positive bias towards patient care outcomes. With regards to some components such as post-operative nausea and vomiting, we identified a particular paucity of evidence in patients undergoing spinal surgery. In these cases, we relied on evidence-based societal recommendations. Full compliance with ERAS protocols has been identified to be an issue in prior studies. Compliance with ERAS pathways has been deemed to be a 5-year survival measure (264). Overall compliance with ERAS protocols has been shown to be associated with better patient reported outcome measures (265).

We have undertaken a number of steps to minimize the underlying meta-biases in this systematic review of complex intervention. We disseminated this protocol through open literature in order to give transparency to our research structure. We assessed the risk of bias in all individual studies. Furthermore, we graded the risk of bias across outcomes (266). As we have identified 22 components of this pathway, some selection bias due to not identifying all eligible studies was possible. Publication bias across studies, where only data published through positive findings are disseminated poses a risk in any systematic review. Detection bias may have arisen due to problems with classification of exposure or outcomes.

## Conclusion

This pathway with an evaluated evidence underpinning each component integrates existing knowledge into practice. Comprehensive evidence based program facilitates institutional perioperative care of spinal surgical patients in the field of ERSS.

## Data Availability

The datasets used and/or analyzed during the current study are available from the corresponding author on reasonable request;

## Declarations

### Ethics approval and consent to participate

Not applicable as this was a systematic review of available literature;

### Consent for publication

Not applicable as this was a systematic review of available literature;

### Competing interests

The authors declare that they have no competing interests.

### Funding

No external funding

### Authors’ contributions

AL: study development, data collection, data editing; first draft manuscript; final version of manuscript; AS: study development, data collection, manuscript editing, final version of manuscript; HL study design, manuscript editing; JR: study design, manuscript editing; CW: study design, manuscript editing;

## Acknowledgements

The authors gratefully acknowledge Mr S. Heaton for his help in data collection.

## Notes

### Competing Interest Statement

The authors have declared no competing interest.

### Clinical Trial

International Register of Systematic Reviews identification number CRD42019135289

### Author Declarations

There was no human data collection, as such the Ethics approval was waived;

